# An open-source dataset of anti-VEGF therapy in diabetic macular oedema patients over four years & their visual outcomes

**DOI:** 10.1101/19009332

**Authors:** Christoph Kern, Dun Jack Fu, Josef Huemer, Livia Faes, Siegfried K. Wagner, Karsten Kortuem, Praveen J. Patel, Konstantinos Balaskas, Robin Hamilton, Dawn A. Sim, Pearse A. Keane

## Abstract

**PURPOSE:** To evaluate visual acuity (VA) outcomes of intravitreal anti-vascular endothelial growth factor (VEGF) in diabetic macular oedema (DMO).

**METHODS:** In this retrospective cohort study, electronic medical records for all patients undergoing intravitreal injections (IVI) in a tertiary referral centre between March 2013 and October 2018 were analysed. Treatment response in terms of visual acuity outcomes were reported for all eyes over a 4-year observation period.

**RESULTS:** Our cohort includes 2616 DMO eyes of 1965 patients over 48 months. Cox proportional hazards modelling identified injection number (hazard ratio [HR] = 1.18), male gender (HR = 1.13), and baseline VA (HR = 1.09) as independent predictors to reach a favorable visual outcome of more than 70 Early Treatment Diabetic Retinopathy Study (ETDRS) letters. Half of our cohort reached 70 letters 1.9 months after starting anti-VEGF therapy. Of those that reached 70 letters, 50% fell below 70 by 14.7 months.

**CONCLUSION:** To date, this is the largest single centre cohort study and over the longest observation period reporting on real-life outcomes of anti-VEGF in DMO. We have made an anonymised version of our dataset available on an open-source data repository as a resource for all clinical researchers globally.

**SYNOPSIS:** Using time-to-event analysis in patients receiving anti-VEGF for DMO: age, baseline visual acuity and injection number are independent predictors of visual outcomes.

## INTRODUCTION

There are currently 21 million patients with diabetic retinopathy (DR) worldwide, which is expected to increase with the projected prevalence of diabetes mellitus (DM) from 415 million in 2015 to 642 million in 2040.[1] The overall risk of diabetic macular oedema (DMO) in patients with DM is currently estimated at 7% (and at 29% after 20 years of disease duration), thus establishing it as the major cause for moderate vision loss in diabetic patients.[2] Randomised controlled trials (RCTs) have demonstrated that intravitreal injections (IVI) with anti-vascular endothelial growth factor (VEGF) agents improve the prognosis of patients with DMO in terms of visual acuity (VA) when following a fixed intervals treatment regimen.[3–6]

In generating the evidence base for optimal patient management, analyses of real-world clinical data have become complements to clinical trials. Evaluation of real-world outcomes ensures continued endorsement of therapeutics by regulators, payers, clinicians, and patients. Compared with clinical trials, real-world studies typically feature a larger sample size and with it, greater heterogeneity amongst its patient cohort and healthcare delivery systems. Such data enable holistic understanding of a therapeutic as they include a more accurate representation of the patient cohorts that receive a therapeutic, how it is used, and the resultant outcome. Notably, treatment conditions in the pivotal anti-VEGF in DMO RCTs are not reflected in real-life clinical settings. A key example is that real-life practice features lower injection frequencies when compared to RCTs.[7–16] This is likely due to different treatment regimens (pro re nata and treat-and-extend) and reduced therapy adherence. Consequently, visual outcome of daily clinical-practice remains unclear.

Here we report on the largest retrospective cohort of DMO patients (2616 eyes of 1965 patients) receiving anti-VEGF therapy and over the longest observation period (48 months) to date. Time-to-event analyses and Cox proportional hazards modelling were carried out to evaluate key positive- (reaching VA of 70 ETDRS letters or greater and remaining above 70) and negative-visual outcomes (VA loss ≥15). An anonymized version of this dataset and our analyses will be made available in an open-source digital repository to increase transparency, accessibility, and permit independent replication of our results. Such availability also enables our data to contribute toward top tier evidence and greater clinical impact.

## METHODS

### STUDY SETTING AND DESIGN

This study is a retrospective cohort study of diabetic patients treated for DMO by anti-VEGF at a tertiary referral centre - Moorfields Eye Hospital NHS Foundation Trust, London, UK. We obtained approval by the Institutional Review Board of the hospital (ROAD17/031) - Audit registration was completed (MEH-233). In this study, we complied with the Declaration of Helsinki and STROBE guidelines for the reporting of cohort studies.[17]

### DATA SOURCE

All clinical information at Moorfields Eye Hospital is recorded within an electronic medical record (EMR) application (OpenEyes Foundation, London, UK). A SQL database (SQL Server Reporting Service, Microsoft Corporation, Richmond, USA) containing all the information from the EMR is in place and regular updates are performed overnight to keep the data warehouse up-to-date. VA is reported in ETDRS letter score. The highest value (independent of measurement method) available at each visit was chosen.

### PARTICIPANTS

A data-warehouse query for patients that received one IVI for DMO (between March 2013 and October 2018) resulted in 3226 unique eyes from 2368 patients. Exclusion criteria were those that: (i) suffered from macular oedema secondary to other conditions than diabetes; (ii) under 18 years old; (iii) received fewer than 3 IVI; (iv) received bevacizumab, dexamethasone intravitreal implant, or fluocinolone acetonide intravitreal implant; leaving 2616 eyes of 1965 patients taken forward for analysis.

### TREATMENT REGIMEN

Patients that were included in this study received anti-VEGF therapy according to the recommended National Institute for Health and Care Excellence (NICE) guidelines at the injection clinic of Moorfields Eye Hospital NHS Foundation Trust, which is approved by the Clinical Audit and Effectiveness Committee (**Supplemental Figure 1**).[18]

**Figure 1.**
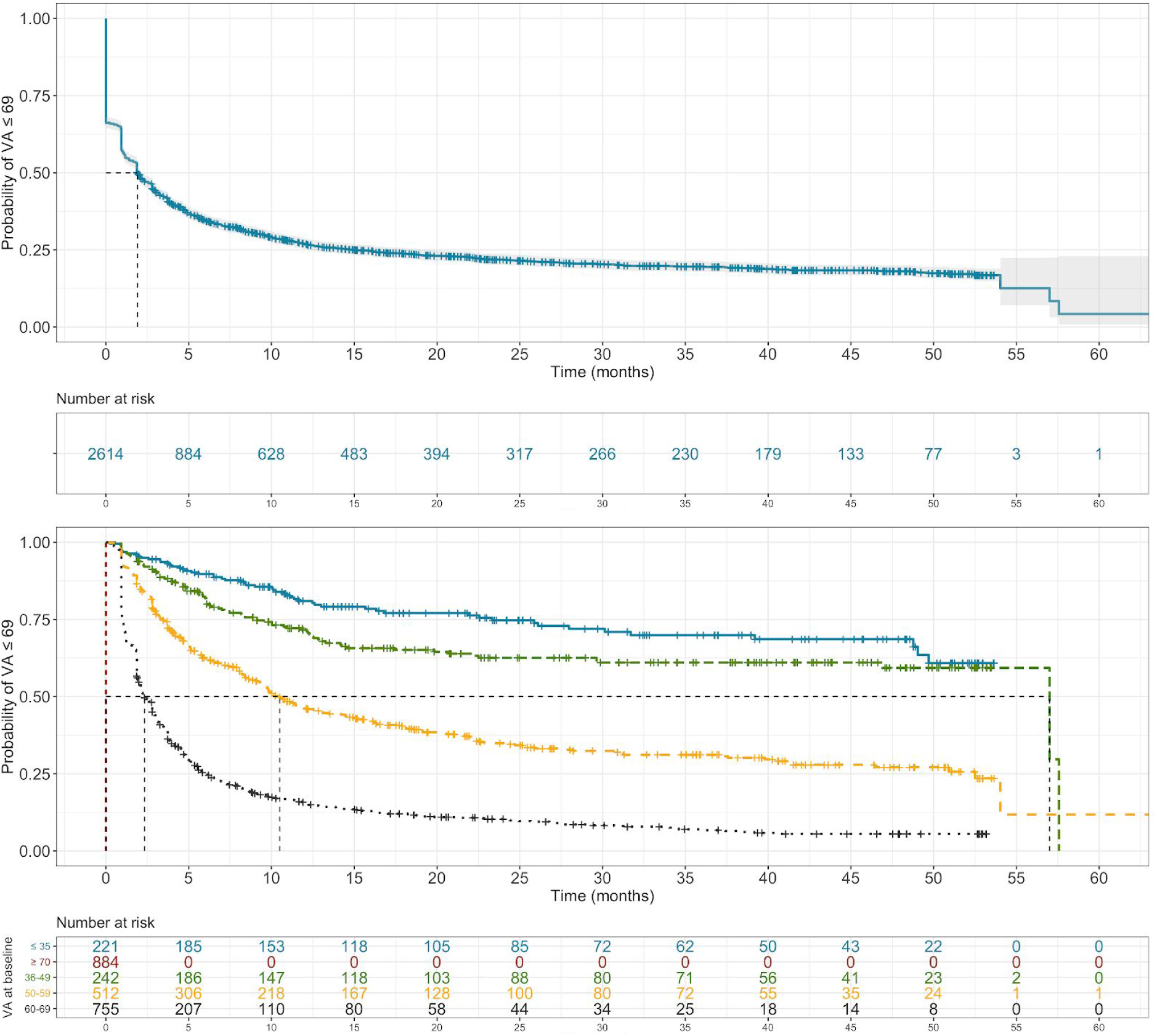
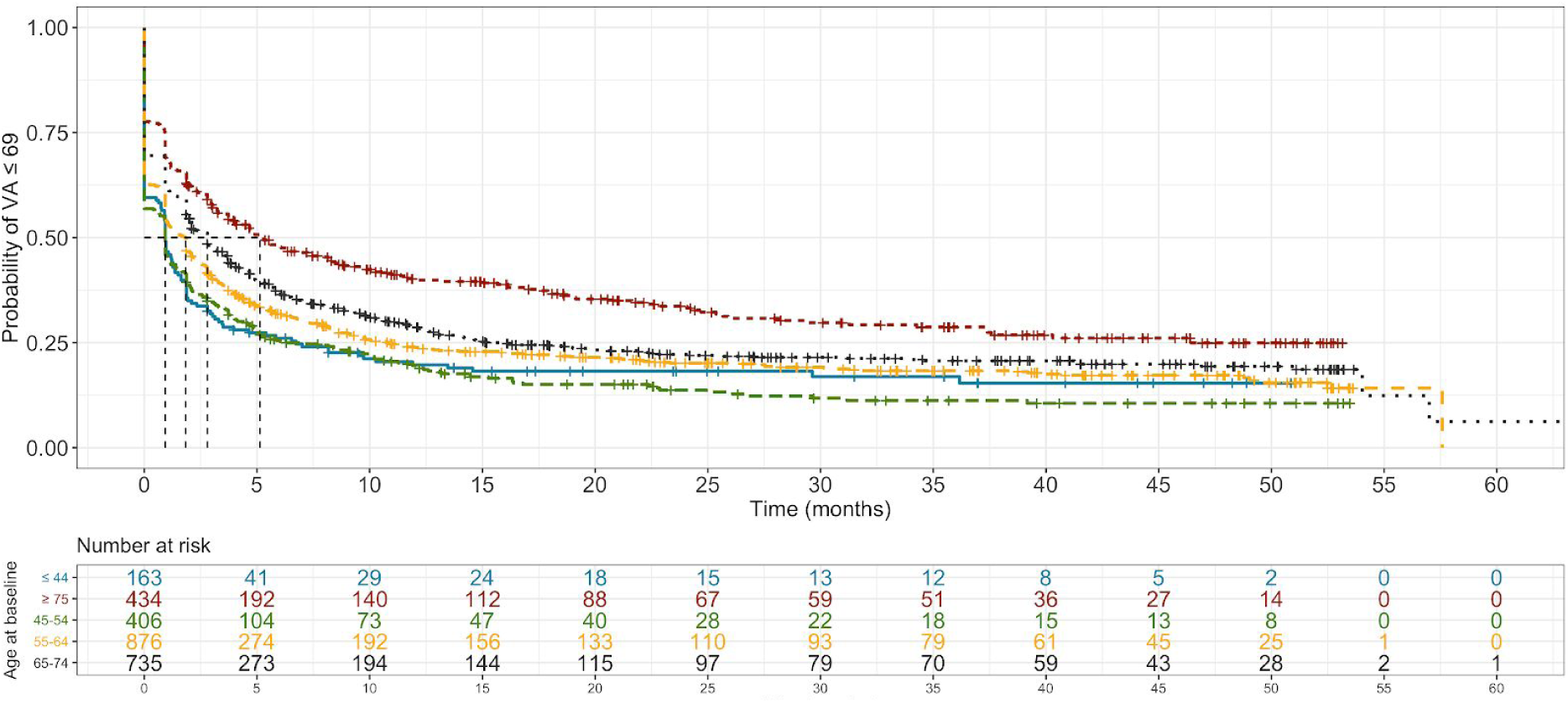
Time-to-event analysis with outcome being visual acuity greater than or equal to 70 ETDRS letters. Time from starting anti-VEGF injections to visual acuity (VA) reaching 70 ETDRS (early treatment diabetic retinopathy study) letters or more was modelled (Top panel). Cohorts were further stratified by statistically significant predictors (**Table 2**): baseline visual acuity (VA; Middle panel) and age at first injection (Bottom panel).

### STUDY OUTCOMES

The primary study outcome is time-to-event analyses (Kaplan-Meier plot) for absolute VA attaining 70 ETDRS letters or above and Cox proportional-hazards modelling to identify predictive covariates. VA was recorded as the best value of the days visit (without correction, glasses or pinhole). Secondary event outcomes include: time to VA less than 70 (≤ 69) and time to VA loss ≥ 15. Time-to-event analyses were employed to obviate the survival bias encountered in traditional visual outcome at given time point metrics (e.g. mean change in VA at year 2), which typically discounts absent patient data. Moreover, by incorporating all the available data leading up to missing values in our models for VA outcomes, they ought to reflect real life more accurately.

**Table 1.** Patient demographics, number of eyes observed and visual acuity outcomes. Eyes were classified to ongoing treatment, observation only, datapoint not available and treatment duration less than time-point.

|  | Baseline | 12 months | 24 months | 36 months | 48 months |
| --- | --- | --- | --- | --- | --- |
| **Patients** | 1964 | 1409 | 1137 | 800 | 479 |
| **Gender (% female:male)** | (40:60) | (41:59) | (41:59) | (40:60) | (42:58) |
| **Age (mean±SD)** | 63±12 | 64±12 | 65±12 | 66±11 | 67±11 |
| **Eyes** | 2614 | 1904 | 1533 | 1051 | 620 |
| **Injections per eye (mean±SD)** |  | 6.4±2.4 | 8.9±4.3 | 11.1±6.0 | 14.0±8.0 |
| **Deaths (%)** | 0 | 1 (0.0005%) | 45 (0.02%) | 92 (0.05%) | 102 (0.05%) |
| **Eyes seen in clinic** | 2614 | 1904 | 1533 | 1051 | 620 |
| - Receiving treatments | 2614 | 1345 | 841 | 509 | 256 |
| - Observation only | 0 | 559 | 692 | 542 | 364 |
| **Eyes not seen in clinic (%)** | 0 | 710 (27%) | 1081 (41%) | 1563 (60%) | 1994 (76%) |
| - Datapoint not available | 0 | 86 | 29 | 15 | 2 |
| - Treatment duration less than time-point | 0 | 624 | 1052 | 1548 | 1992 |
| **Mean visual acuity** | 61.0 ± 15.3 | 66.2 ± 15.5 | 65.8 ± 15.8 | 64.1± 17.0 | 61.8 ± 18.2 |
| (ETDRS letters ± SD) | | | | | |
| Mean change in visual acuity (ETDRS letters) | - | 5.2 ± 12.8 | 4.8 ± 13.5 | 3.4 ± 15.7 | 2.5 ± 17.3 |

**Table 2.** Hazard ratios derived by Cox-models for certain events: Visual acuity greater or equal than 70 ETDRS letter, Visual acuity dropping below 70 letters in patients that reached VA above 70 ETDRS letters under therapy and vision loss of more than 15 ETDRS letters

|  | Hazard Ratio |  | Lower |  | Higher |  | p-value |
| --- | --- | --- | --- | --- | --- | --- | --- |
| **Cox model for visual acuity $\geq 70$ ETDRS letters** |  |  |  |  |  |  |  |
| Gender - Male | 1.13 | ( | 1.03 | - | 1.24 | ) | <0.001 |
| Age @ baseline | 0.99 | ( | 0.98 | - | 0.99 | ) | <0.001 |
| VA @ baseline | 1.09 | ( | 1.09 | - | 1.1 | ) | <0.001 |
| Number of injections | 1.18 | ( | 1.14 | - | 1.22 | ) | <0.001 |
| **Cox model for visual acuity $\leq 69$ in those that have reached VA $\geq 70$** |  |  |  |  |  |  |  |
| Gender - Male | 0.92 | ( | 0.82 | - | 1.04 | ) | 0.172 |
| Age @ baseline | 1.01 | ( | 1.01 | - | 1.02 | ) | <0.001 |
| VA @ baseline | 0.97 | ( | 0.96 | - | 0.97 | ) | <0.001 |
| Number of injections | 0.96 | ( | 0.93 | - | 0.99 | ) | <0.01 |
| **Cox model for visual acuity loss of more than 15 ETDRS letters** |  |  |  |  |  |  |  |
| Gender - Male | 0.88 | ( | 0.74 | - | 1.04 | ) | 0.130 |
| Age @ baseline | 1 | ( | 0.99 | - | 1.01 | ) | 0.999 |
| VA @ baseline | 1.01 | ( | 1 | - | 1.01 | ) | <0.05 |
| Number of injections | 0.98 | ( | 0.96 | - | 1 | ) | <0.05 |

To enable comparison with previously published functional outcomes, we also carried out : (i) mean VA and change in VA per study eye as compared to baseline in ETDRS letters; (ii) proportion of eyes with change in VA being < 10 ETDRS letters and ≥ 10 ETDRS letters; and (iii) proportion of eyes with ≥ 15 ETDRS letters gain or loss. For these analyses, selected observation time-points and their definitions were as follows: baseline (date of first intravitreal injection); one year (12 months; 365 days ± 90 days); two years (24 months; 720 ± 150 days); three years (36 months; 1095 ± 150 days); four years (48 months; 1460 ± 150 days).

### STATISTICAL METHODS

For statistical analyses, all EMR data was handled in R.[19] Time to each of the visual outcomes were visualised with Kaplan-Meier time-event plots. Cox proportional hazards regression models were also carried out to evaluate the effects of demography (gender, ethnicity), clinical features at baseline (age and VA), and IVI (included as time-dependent covariates). Distribution of data was tested by the Shapiro-Wilk normality test. Means of non-parametric groups were compared using Wilcoxon Signed-rank, Wilcoxon Rank-sum, and Kruskal-Wallis tests as appropriate. For more than two groups, multiple pairwise-analyses was carried out with the Wilcoxon Rank-sum test. Calculated means in text and figures are expressed with ± error margin corresponding to the standard deviation, unless otherwise specified. A p-value < 0.05 was considered statistically significant.

### DATA SHARING AGREEMENT

An anonymised version of the dataset as well as the code used for analysis is available in the open source digital repository Dryad - https://doi.org/10.5061/dryad.pzgmsbcfw. Depersonalisation was carried out through hash function anonymisation of patient identification numbers, and replacement of appointment dates with follow-up days to baseline. Approval of adequate depersonalisation was obtained by Moorfields Information Governance.

## RESULTS

### PATIENT CHARACTERISTICS

Our cohort comprised 2616 eyes of 1965 patients who initiated and completed a loading course of anti-VEGF therapy within the 4-year observation period (**Supplemental Figure 2**). We compiled a dataset that includes demographic information (**Table 1**) and all available VA-values and IVIs from baseline (initiating of therapy) through to end of observation period.

**Figure 2.**
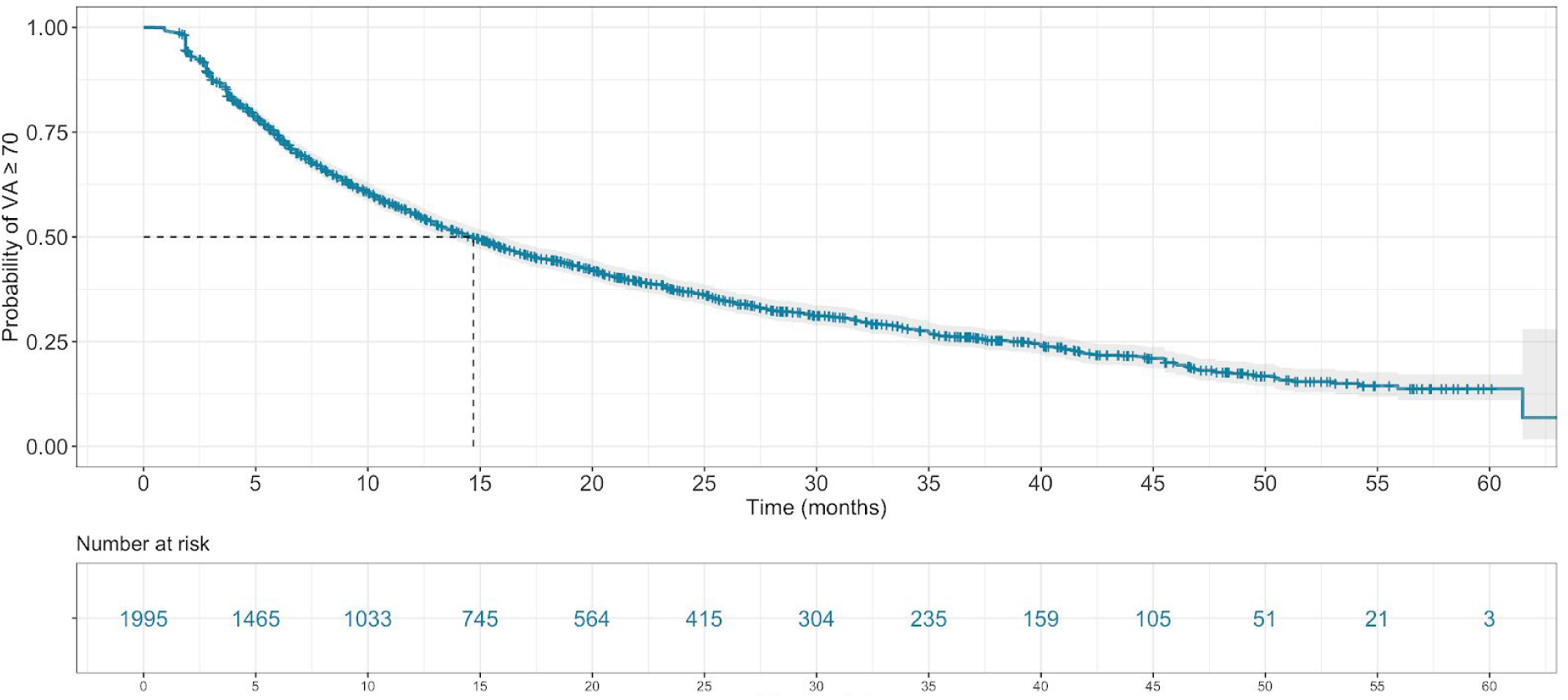
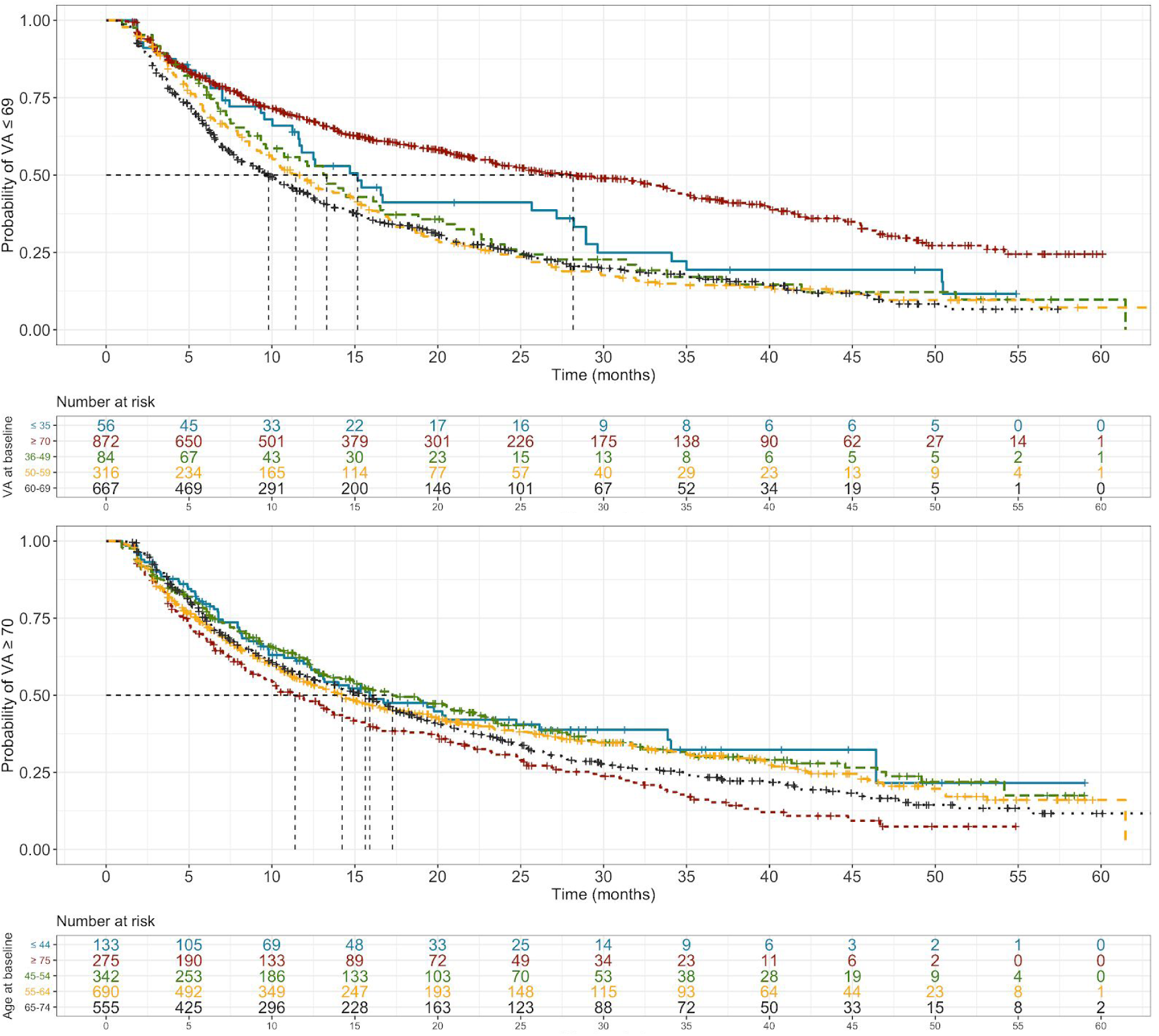
Time-to-event analysis with outcome being visual acuity less than 70 ETDRS letters. All patients attaining visual acuity (VA) equal to or greater than 70 ETDRS (early treatment diabetic retinopathy study) letters in the observation period were taken forward for analysis. Date of reaching this threshold was considering baseline, with time to VA falling below 70 modelled (Top panel). Cohorts were further stratified by statistically significant predictors (**Table 2**): baseline visual acuity (VA; Middle panel) and age at first injection (Bottom panel). Of note, the sub-cohort of patients that began with VA at or over 70 (Red line; Middle panel) were included in sub-stratified by baseline VA and featured median event time almost two-fold greater than the other sub-cohorts.

At baseline, the mean VA was 61.0 ± 15.3 ETDRS letters. For each of the annual timepoints, we saw an increasing number of eyes with missing VA values. This was largely due to the treatment duration for a given patient being shorter than the timepoint itself, leaving few due to loss-to-follow-up. For instance, of the 1995 eyes without data at the 4-year timepoint, only 2 were lost-to-follow-up with the remainder having a treatment duration less than 4 years.

### MEAN VISUAL ACUITY TRENDS ARE SIMILAR TO EXISTING LITERATURE

Trends in VA were compared to previously published studies. Mean VA changes and IVIs were comparable to retrospective cohort studies at one- and two-year timepoints (**Supplemental Table 1**). Data for comparison of three- and four-year timepoints do not yet exist. A change in VA of 10 and 15 letters are frequently considered when evaluating outcomes in DMO patients receiving anti-VEGF therapy. [13,15,20] Also in keeping with reported retrospective studies, the proportion of eyes in this cohort gaining ≥ 10 letters was greater than 30% and exceeded those who lost ≥ 10 letters at each of the annual time-points (**Supplemental Figure 3a**).[13,15,20] A similar trend was observed when considering the proportion of eyes which gained ≥ 15 letters, with 19.3%, 20.5%, 18.9%, and 21.3% of eyes observed at the annual time-points spanning 1 to 4 years following baseline **(Supplemental Figure 3b**).

**Figure 3.**
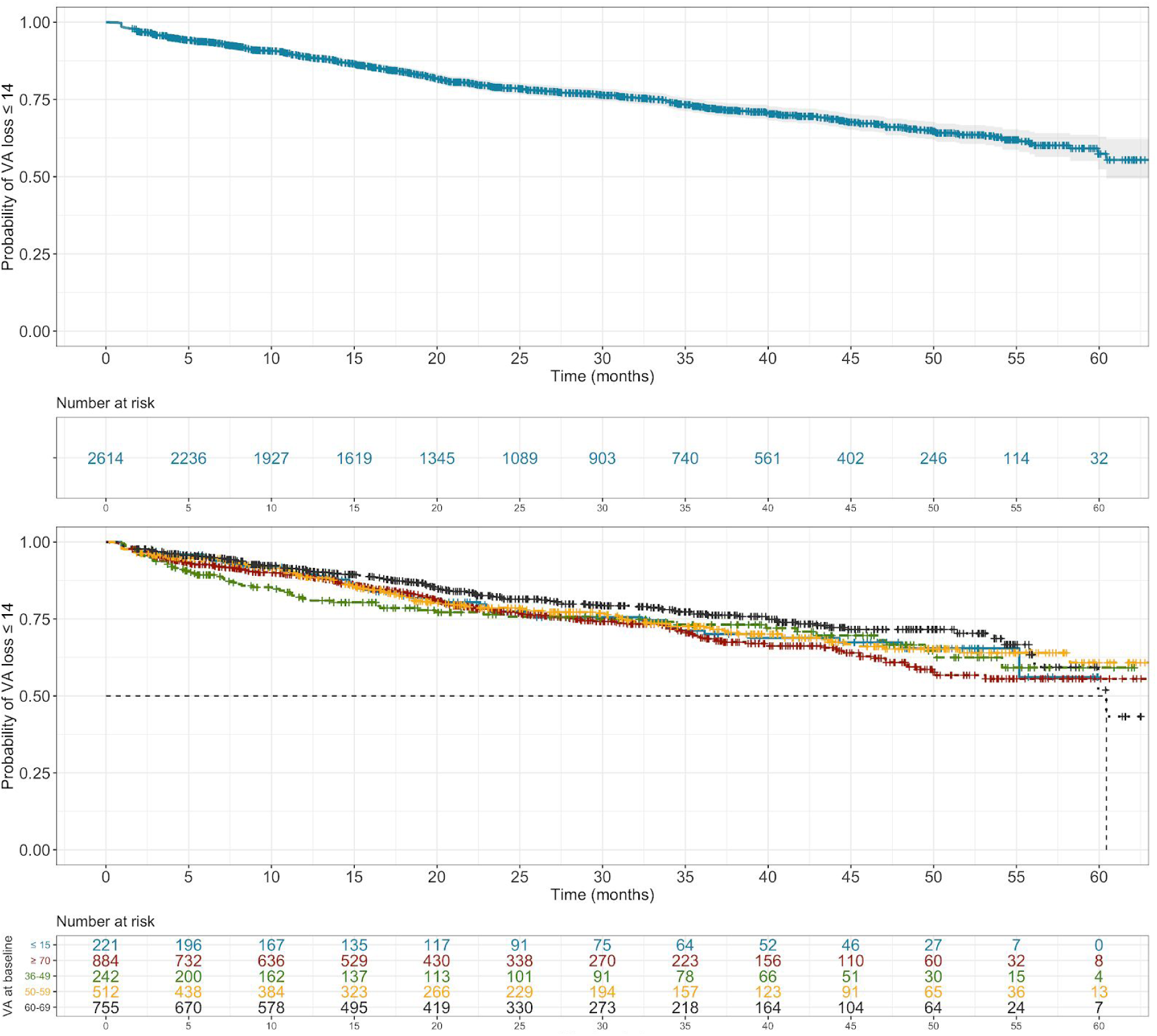
Time-to-event analysis with outcome being visual acuity loss of 15 or greater. Date of injection 1 was considered baseline and time to visual acuity (VA) of 15 ETDRS letters or more was modelled (Top panel). Cohort was further stratified by the statistically significant predictor (**Table 2**) baseline VA (Bottom panel).

### PREDICTIVE FACTORS FOR POSITIVE VISUAL OUTCOMES

Absolute VA ≥ 70 ETDRS letters is commonly used to measure positive visual outcomes in a clinical setting. Kaplan-Meier modelling of our cohort data suggests that 50% of DMO eyes are likely to reach VA ≥ 70 at 1.9 months after starting anti-VEGF therapy and over 75% after a year (**Figure 1a**). To identify predictive variables for VA ≥ 70 in our cohort, we used Cox proportional-hazards modelling to relate VA outcomes to clinical time-dependent (anti-VEGF injections) and time-independent (gender, age at baseline, VA at baseline) covariates. This suggests that number of IVI, being male and - to a greater extent - VA at baseline are positively associated with VA ≥ 70 (**Table 2**).

Indeed, the time point at which 50% of people are likely to reach VA ≥ 70 is much sooner in those with higher baseline VA of 60-69 (2.3 months) than with a lower baseline VA of 36-49 (57.0 months) (**Figure 1b**). In contrast, age at baseline correlates inversely with VA ≥ 70 (**Table 2**) and with a smaller impact on event probability. A small, but statistically significant, difference in median event time is even seen between patients at either extremes of age - 0.9 months in those ≤ 44 years of age compared with 5.1 months in those ≥ 75 (**Figure 1c**).

### INTERROGATING DURATION TO VISION LOSS

The majority of our cohort attained the positive clinical outcome of VA ≥ 70 during the 48 month observation period (1995 of 2616 eyes). We subsequently interrogated the duration for which the VA remained at or above 70 in this sub-cohort by modelling time between attaining VA ≥ 70 to falling below 70 (≤ 69 letters). Of patients that reach VA ≥ 70, 50% are likely to fall below 70 at 14.7 months (**Figure 2a**). Statistically significant predictive variables for falling below 70 after having attained VA ≥ 70 are VA at baseline (**Figure 2b**), age at baseline (**Figure 2c**), and injection number (**Table 2**). Interestingly, gender does not contribute significantly to VA falling below 70 as it does for reaching 70.

### PREDICTIVE VARIABLES FOR NEGATIVE VISUAL OUTCOMES

A significant loss in vision has often been defined as loss of 15 ETDRS letters in the research of macular diseases as a synonym for a three lines decline on Snellen chart. It is also recommended by the FDA as a relevant outcome for studies addressing macular diseases.[21] We observed a trend of male sex reducing the risk of losing more than 15 ETDRS letters during therapy. Despite the strong effect of HR of 0.88 this predictive factor was not statistically significant. Independent predictors for losing more than 15 letters were a low VA at baseline as well as a low number of injections (**Table 2**). As expected, age at baseline did not contribute as a predictor for unfavorable vision loss. [22]

## DISCUSSION

### MAIN FINDINGS

Here we report on the largest retrospective cohort of DMO patients (2616 eyes of 1965 patients) receiving anti-VEGF therapy and over the longest observation period (4 years) to date. Reaching VA of 70 ETDRS letters or greater is an indicator of patient independence. Indeed, it is: (i) used as the mark for low vision alongside visual field loss and loss of contrast sensitivity; [23,24] (ii) the threshold for driving; [25] (iii) the minimum VA required to read the small print; [26] and therefore chosen as key positive VA outcome. The majority of our cohort achieved VA ≥ 70 (76%) over the observation period and were likely do so shortly after initiating anti-VEGF therapy (median survival at 1.9 months; **Figure 1**). However, it is unknown for how long a given eye can expect to remain above this critical level. Our analyses demonstrate the median survival of this sub-cohort to remain at or above 70 is 14.7 months (**Figure 2**). That is, the VA in 50% of eyes will fall below 70 ETDRS letters 14.7 months after reaching ≥ 70. Baseline VA, baseline age, and IVI number are predictive covariates for VA reaching and remaining above 70 (**Table 2**). An interpretation of these data is that early recognition of DMO indicated for treatment can increase the probability of positive visual outcomes; including VA reaching 70 ETDRS letters and extending the duration one remains above it. [4,5,27]

### VISUAL OUTCOMES IN RCTs

Demography and trends in VA of our cohort are consistent to those in similar real-world data reports in DMO (**Supplemental Table 1**). However, the annual mean VA changes observed in our cohort were less than those reported in the RCTs that led to approval of the anti-VEGF treatment (**Table 1**). [3–5] It has been postulated that these differences in IVI regimen and number account for this. For instance, the mean IVIs delivered over 24 months in the RISE (20.9) and RIDE (21.9) studies [4] are two-fold greater than in any study reporting real-world data (**Supplemental Table 1**), including ours (8.9; **Table 1**). Our cohort analyses support this as they demonstrate IVI as a positive predictive covariate of VA reaching VA ≥ 70 (HR 1.18 [95%CI 1.14 - 1.22]) and protective for negative VA outcomes - VA falling below 70 (HR 0.96 [95%CI 0.93 - 0.99]) and VA loss ≥ 15 (HR 0.98 [95%CI 0.96 - 1.00]) (**Table 2**). It is important to note that our model represents the Moorfields treatment protocol and incorporates IVIs as a time-dependent covariate. As such, patients that receive IVIs could signify greater disease severity. It is notable that IVIs are statistically significant predictors of positive visual outcomes in spite of this.

Another potential reason for the discrepancy in annual mean VA change between RCTs and our study is a difference in baseline VA. Our cohort features a greater mean baseline VA than the RCTs (**Table 1**). It is established that baseline VA is inversely correlated with mean VA changes at 1 and 2 years in DMO patients receiving anti-VEGF therapy [28]; as is evident from the broad range of mean baseline VAs (48 to 63) featured in real-world studies (**Supplemental Table 1**. Interestingly, our model identifies baseline VA as a protective factor for positive outcomes: VA reaching 70 or more ETDRS letters (HR 1.09 [95% CI 1.09-1.1]) and remaining above 70 (HR=0.97; [0.96-0.97]) (**Table 2**). That is, although those with a low baseline VA may exhibit a greater VA increase at 1 and 2 years, they are less likely to reach a positive visual outcome than those with a higher baseline VA.

### ADDRESSING MISSING DATA

Loss-to-follow-up (LTFU) is a general and challenging limitation in retrospective cohort data when evaluating outcomes at given time points. This is also the case in studies looking at anti-VEGF in DMO. LTFU rates reported in literature are as high as 13%, 31%, 48% and 65% after 1, 2, 3 and 4 years, respectively; and even up to 95% after 2 years in multicentre analysis. [8,14] Our data featured comparable missing data at key time points i.e. 27%, 41%, 60% and 76% at 1, 2, 3, and 4 years, respectively (**Table 1**). We therefore scrutinised the underlying reason for missing data by considering the treatment duration for each eye. As expected, we saw that the majority of missing data (>85% at each time point) were due to treatment duration being shorter than the time point (**Table 1**). For example, a patient with DMO initiating treatment in 2018 will not have a value any of the annual time points and it would be inappropriate to label them as LTFU. Through this consideration alone, our dataset features the lowest LTFU rate reported amongst similar studies.

We also considered whether values were missing due to patients being deceased and is the first of such studies to do so. Interestingly, data values from 163 eyes of 102 patients were absent due to death over a 4 year observation period which extrapolates to 30.6 deaths per 1000 person-years. This is higher than what has been previously reported in DM (26 deaths per 1000 person-years). [29] This correlation may be explained over the amount of comorbidities besides diabetes that are present in the UK population. [30] Moreover, one would expect our study cohort (patients receiving hospital treatment for complications of DM) to be at higher risk of comorbidities than the general DM population.

### LIMITATIONS OF OUR DATA

Our dataset attempts to account for missing data and consequently features the lowest LTFU rate amongst similar studies. Although this is the case, it is important to note that missing values remain and these would be ignored in traditional time point analyses e.g. mean change in VA at year 2. Here it would be controversial to assume that patients with absent data at a given time point (death or otherwise) are a random selection of those that initiate treatment. As such, survival bias would feature prominently if mean values at given time points were generalised to all DMO patients that undergo anti-VEGF therapy. Accordingly, we have employed time-to-event analyses as they use all data up until the point that someone has an event or no longer followed up. Moreover, time-independent and -dependent covariates were incorporated to enable adjustment for age, baseline VA, gender, and injections.

In patients with bilateral DMO involvement and receiving IVIs, both eyes were included in our analyses. This is a possible confounder as it could lead to multiple-testing. However, this could be addressed by other workgroups using our open-source dataset.

### FUTURE IMPLICATIONS

To date, this is the largest single centre cohort study reporting on real-life outcomes of anti-VEGF in DMO. There are several similar real-world data reports in DMO (**Supplemental Table 1**). Of these, our cohort size is larger than six of them combined. Comparable cohort size have been reported by Egan et al. (2017); a UK wide multicentre study with 19 attending centres and thus various distinct treatment regimes, as opposed to a single protocol in our single centre study. [14] Our inclusion criteria are broader than in the pivotal RCTs, including patients with higher and lower baseline VA, thus granting a more realistic account of DMO treatment outcomes.

A key output of this study is to make an anonymised version of our dataset available on an open-source data repository as a resource for all clinical researchers globally. This is with the aim of optimising the capacity of our data to positively impact clinical research and outcomes. By including data beyond the two year time horizon, we hope to expand current insight into long-term visual outcomes by enabling comparison and meta-analysis with future data that also report beyond two years. There is an increasing call for research that is transparent and that can have its key findings reproduced by its audience.[31,32] This is of particular concern in clinical research predicated on digitalisation of healthcare environments, as collective progress can be hampered by the absence of detailed methodology and data sharing. To further address this, we have published a step-by-step guide written in open source code R of how we performed our statistical analyses. This code can be directly applied to the published database to replicate all figures and values in this study.

## Conclusions

Analyses of our cohort reveal that the majority reach 70 ETDRS letters or more whilst receiving their initial loading dose. Of these, the median survival for remaining above 70 is 14.7 months. Furthermore, we demonstrate that age, baseline VA, and injection number are independent predictors of visual outcomes - suggesting that earlier diagnosis and treatment of DMO could increase the likelihood of positive outcomes. Lastly, this is the largest retrospective cohort study of anti-VEGF in DMO over the longest observation period to date and we have made this dataset available.

## Data Availability

An anonymised version of the dataset as well as the code used for analysis is available in the open source digital repository Dryad.

https://doi.org/10.5061/dryad.pzgmsbcfw

## SUPPLEMENTAL MATERIALS

**Supplemental Table 1.**
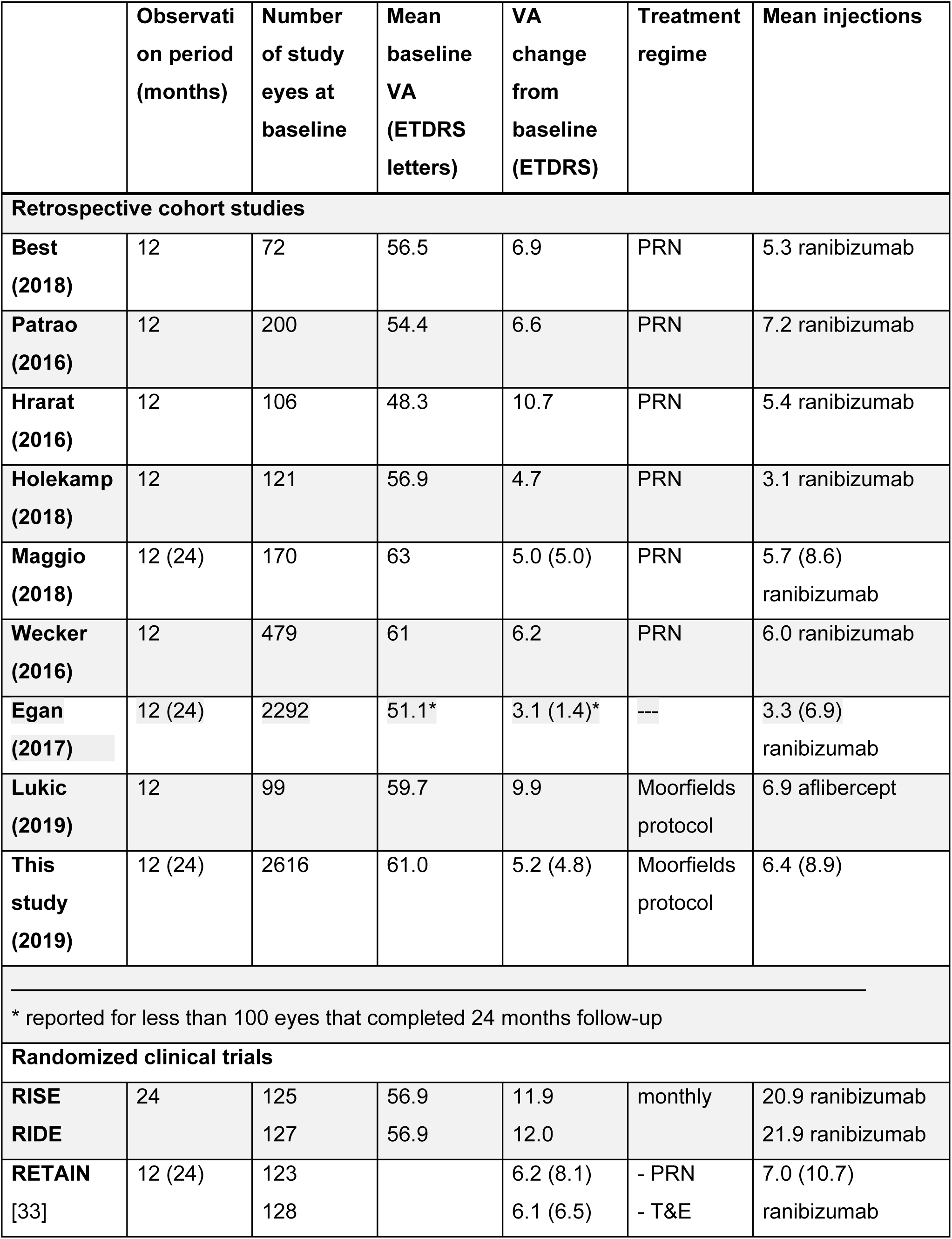
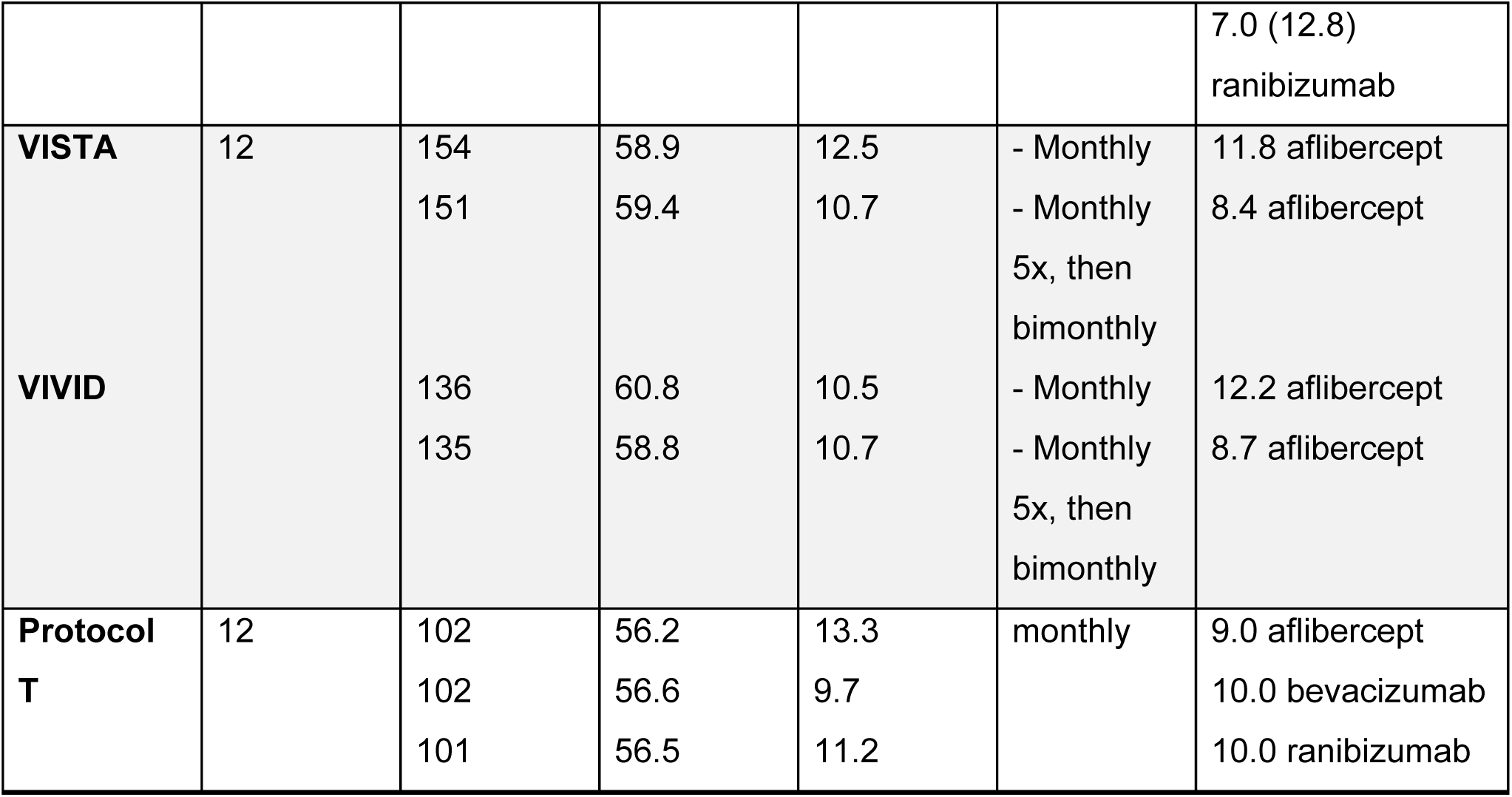
Visual acuity outcomes reported in change from baseline for selected real-life studies and randomized clinical landmark trials, including treatment regimes, delivered drugs, observation period, mean number of injections and number of eyes observed.

**Supplemental Figure 1.**
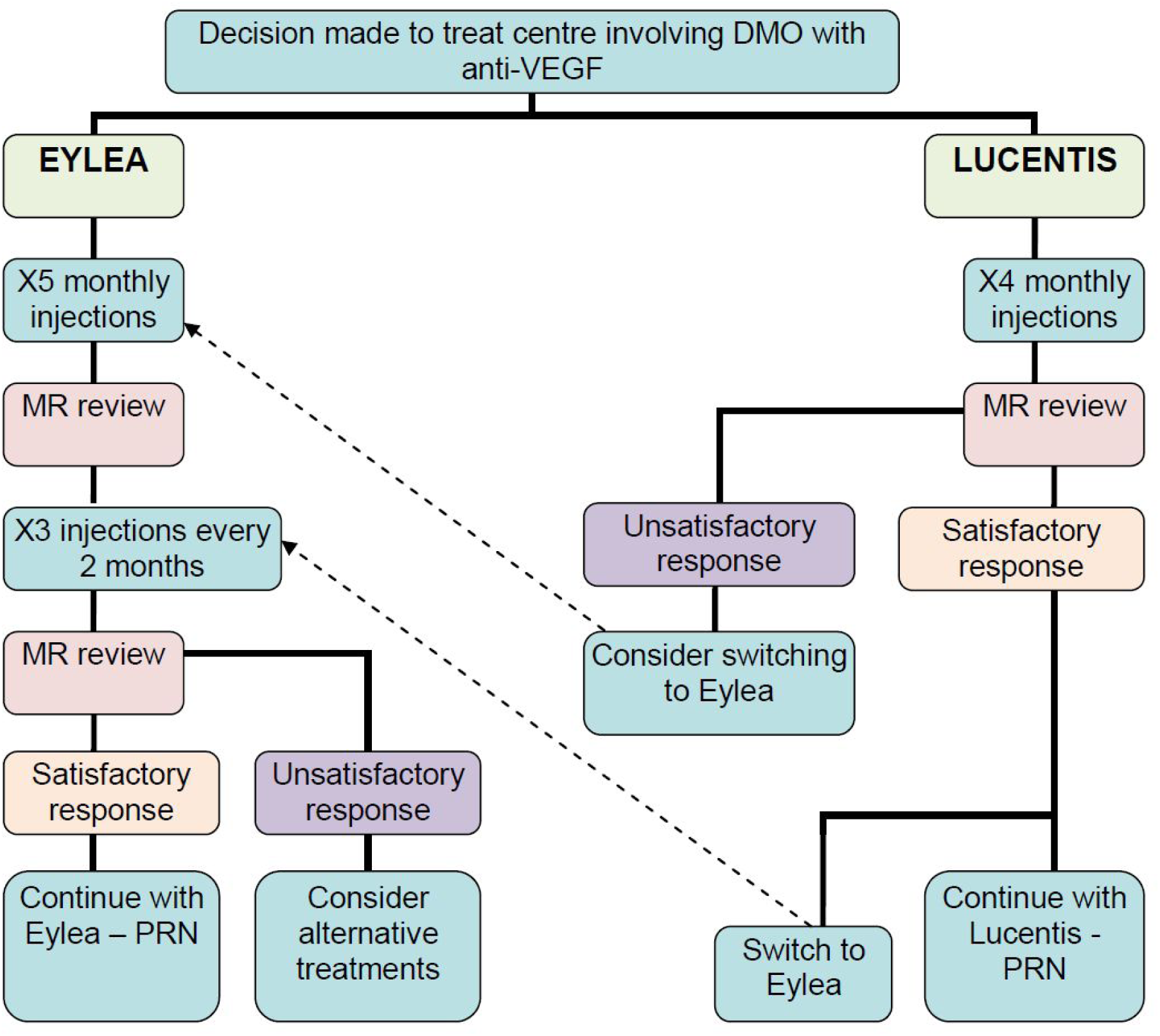
Treatment flow chart for Anti-VEGF treatment of patients with NICE eligible diabetic macular oedema at Moorfields Eye Hospital. Derived from “Protocol for Clinicians working in the Diabetic Macular Oedema (DMO) Injection Clinic” (Version 1.0); approved and ratified by the Clinical Audit and Effectiveness Committee

**Supplemental Figure 2.**
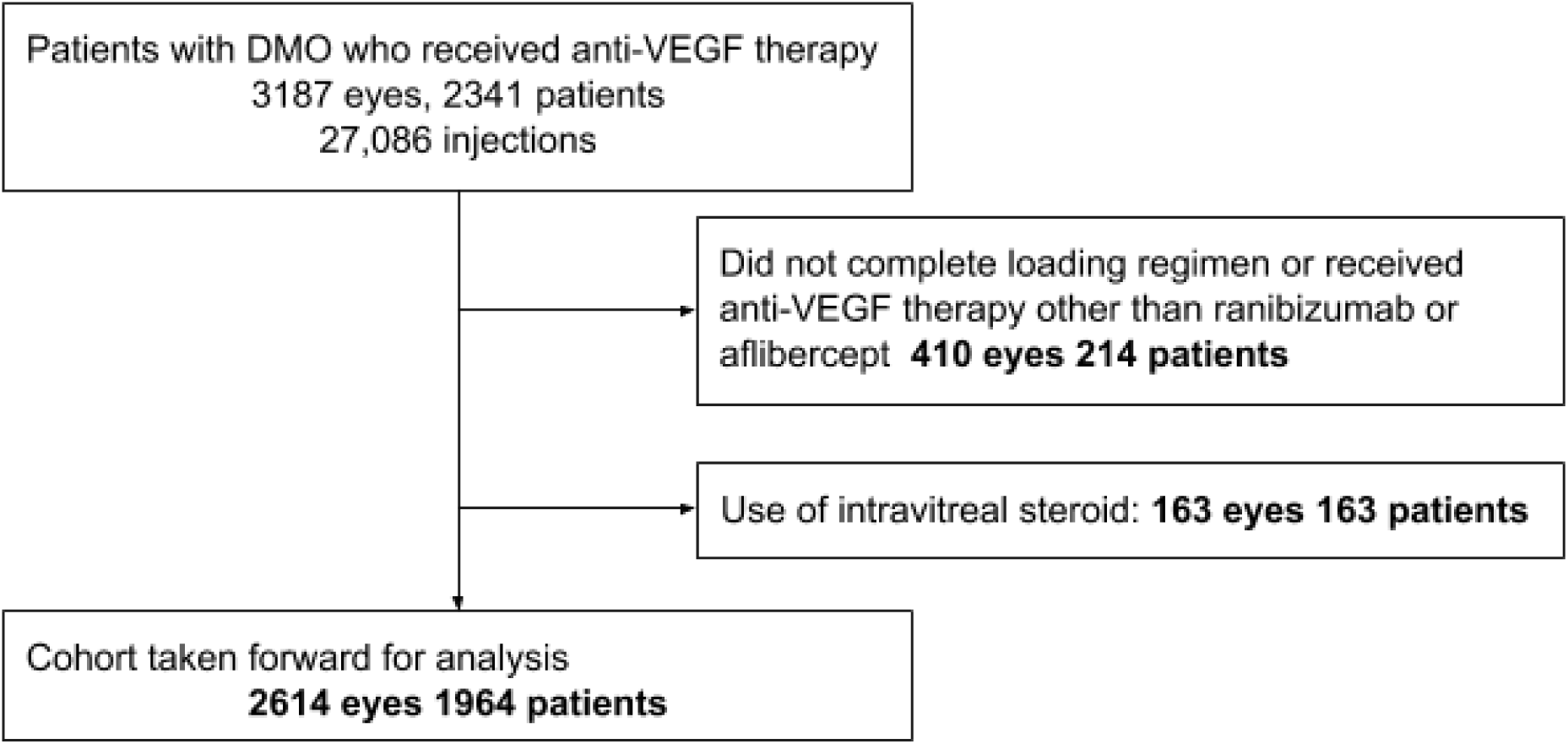
Patient cohort after application of inclusion and exclusion criteria.

**Supplemental Figure 3.**
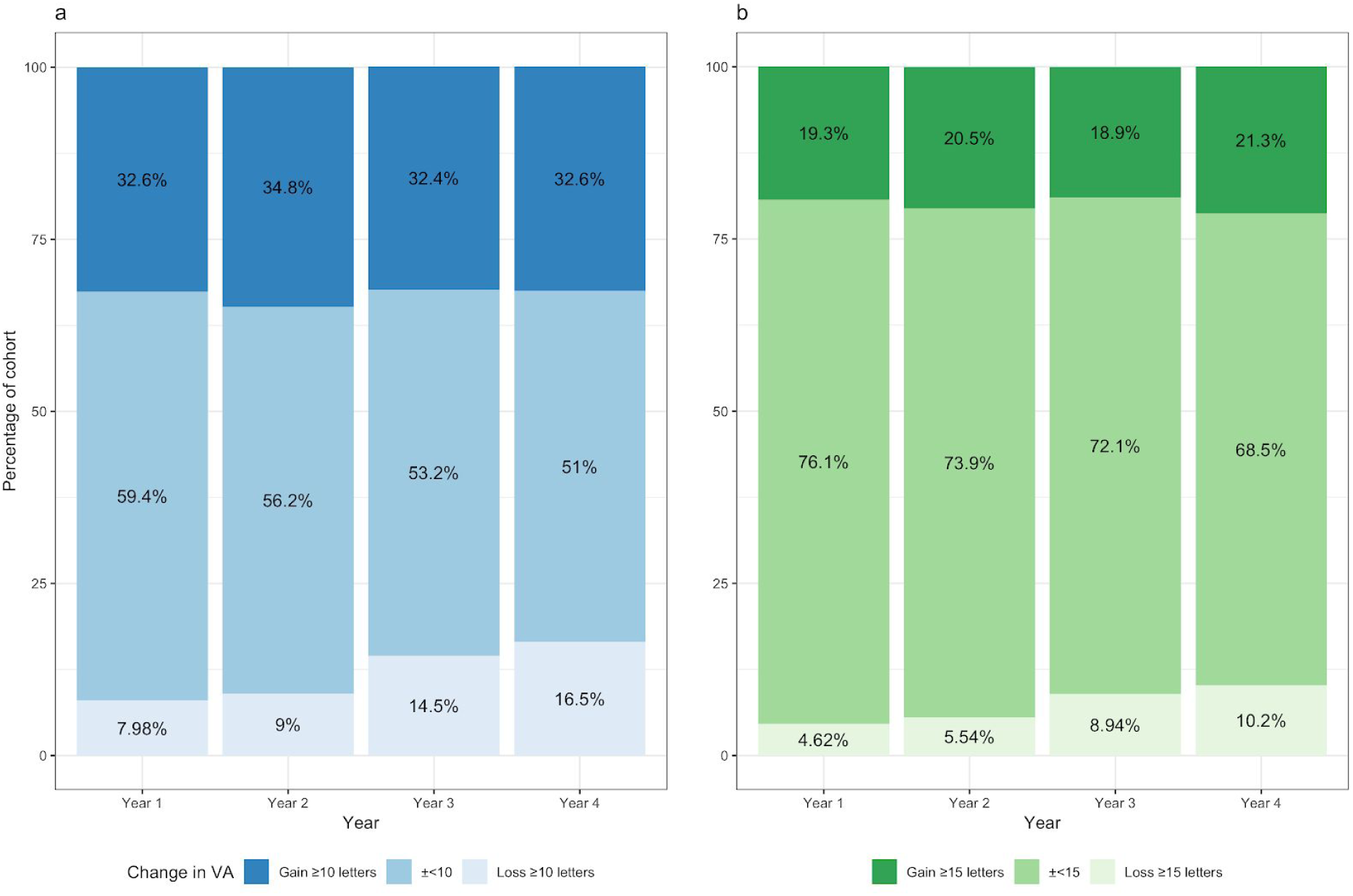
Annual visual outcomes with a change greater than or within 10 and 15 ETDRS (early treatment diabetic retinopathy study) letters. Proportion of eyes (expressed as percentage) with a change in visual acuity within or greater than (a; blue) 10 and (b; red) 15 ETDRS letters. Proportion of eyes that lost ≥ 10 and ≥ 15 letters progressively increased annually. Conversely, eyes that gained or lost letters < 10 and < 15 decreased with each year.

